# UNVEILING URBAN AIR QUALITY SEASONAL DYNAMICS IN SAN MIGUEL DE TUCUMÁN, ARGENTINA, BY SCANNING ELECTRON MICROSCOPY

**DOI:** 10.1101/2025.06.23.25330139

**Authors:** Enzo Rubén Marcial, Alexander Aldo Santucho Cainzo, Facundo Reynoso Posse, Rodrigo Gastón Gibilisco, Aida Ben Altabef, Diego Hernando Corregidor Carrió, Virginia Helena Albarracín

## Abstract

Scanning Electron Microscopy (SEM) coupled with Energy Dispersive X-ray Spectroscopy (EDS) was used to analyze particulate matter (PM) in San Miguel de Tucumán, Argentina, a densely populated urban area influenced by agricultural and industrial pollution. This study investigates PM concentration and composition during sugarcane harvest and non-harvest periods (2019-2020), accidentally coinciding with the COVID-19 lockdowns. SEM-EDS enabled the classification of PM by size (Dust, PM_10_, and PM_2.5_) and by composition (Terrigenous PM and Carbonaceous PM). Higher PM concentrations were detected during harvest periods, with a notable increase in carbon-rich PM_2.5_ (PMC_2.5_) in 2019 compared to 2020. Unexpectedly, this variation was primarily attributed to reduced population mobility during the 2020 lockdowns, rather than changes in industrial activity. Satellite data on nitrogen oxides (NOx) supported these findings, revealing emission patterns consistent with vehicular activity. Overall, this study demonstrates the effectiveness of SEM-EDS in characterizing air pollutants and highlights the critical influence of population mobility on urban air quality. The findings support the implementation of targeted air pollution control strategies - particularly during harvest seasons- to mitigate both health and environmental impacts.

## 1. Introduction

Air pollution poses a major threat to urban environments, particularly in regions where agricultural and industrial activities coexist. Particulate matter (PM), a complex mixture of solid particles and liquid droplets suspended in the air, is a key contributor to degraded air quality, with significant health and environmental consequences (Ministry of Environment and Sustainable Development, 2021; World Health Organization, 2021). The city of San Miguel de Tucumán, Argentina, exemplifies these challenges, being a densely populated urban center surrounded by intensive agricultural operations, notably sugarcane farming, which substantially influences seasonal air pollution patterns.

PM is commonly classified according to aerodynamic diameter into Dust (>10 µm), PM_10_ (<10 µm), and PM_2.5_ (<2.5 µm), each posing specific health risks (Pakbin et al., 2010). Exposure to PM has been associated with a wide range of diseases, including respiratory, cardiovascular, and neurological disorders, as well as increased mortality (Correia et al., 2013; World Health Organization, 2021). In Tucumán, seasonal practices such as sugarcane field burning—despite being legally prohibited by Provincial Law No. 6253, Art. 38—continue to exacerbate air quality problems, contributing to peaks in respiratory and ocular conditions (Altieri et al., 2018; Jordán and Flores, 2018).

PM originates from diverse sources, including industrial activities, vehicular emissions, biomass burning, and agricultural residues. Key components of PM include sulfates, nitrates, ammonia, sodium chloride, mineral dust, ashes, and carbonaceous particles. Carbonaceous particles, forming 20–45% of fine particles, are primarily released through combustion processes, such as biomass and agricultural residue burning (Ji et al., 2016; Dinoi et al., 2017). These particles significantly impair air quality, not only by increasing light absorption in the atmosphere but also by reducing visibility (Zhou et al., 2012; Bisht et al., 2016; Lee et al., 2017), which is one of the most perceptible consequences of atmospheric pollution. The use of organic tracers has identified primary sources of elemental carbon, including coal combustion, vehicle exhaust, biomass burning, and vegetative detritus (Li et al., 2013). In Tucumán, the seasonal burning of sugarcane fields intensifies PM concentrations, complicating air quality management efforts. Although this practice is prohibited by Provincial Law No. 6253, Art. 38, enforcement remains limited, and comprehensive regulations to mitigate air pollution are lacking.

Current air quality monitoring systems in Tucumán, and worldwide, are constrained by technological and methodological limitations, with yet scarce development of innovative, cost-effective analytical techniques. Traditional methods such as gravimetric analysis (Almeida et al., 2006; Chatoutsidou and Lizariadis, 2022) and optical particle counters (Wang et al., 2021, Mills et al., 2024) are widely used but often lack the precision needed to determine the chemical composition of PM or require expensive, specialized equipment. Scanning Electron Microscopy (SEM) coupled with Energy-Dispersive X-ray Spectroscopy (EDS) offers a robust alternative for PM characterization, providing high-resolution insights into particle size, morphology, and chemical composition, this way, it is possible to identify individual particles originating from specific sources such as coal mines, road wear, brakes and wheels, sea salt, soil, and industries(Casuccio et al., 2004;Fanizza et al., 2018; Rausch et al., 2022; Li et al., 2023; Zhao et al., 2023; Aubourg et al., 2024). Despite its potential, SEM-EDS remains an underexploited tool in the context of regional air quality studies.

In this context, the present study explores how seasonal agricultural practices, coupled with variations in human activity, influence the concentration, size distribution, and chemical composition of PM in an urban environment. By applying SEM-EDS to samples collected across harvest and non-harvest seasons—and coinciding with the exceptional conditions imposed by COVID-19 lockdowns—this work aims to elucidate the relative contributions of natural and anthropogenic factors to particulate pollution in San Miguel de Tucumán. Ultimately, the findings are intended to support the development of targeted air quality management strategies in urban areas impacted by agricultural activities.

## 2. Methods

### 2.1. Sampling Design and Collection

Samples were collected at the Quinta Agronómica campus of the National University of Tucumán, located in central-southern San Miguel de Tucumán. An open field was chosen to minimize contamination from nearby trees and buildings.

The sampling device used adhesive carbon tape—which retains settled particles—mounted on an aluminum stub and fixed to a polystyrene base for stability and easy removal (Figure 1). It was placed at a height of 2 meters and stored in airtight glass containers to prevent contamination during transport.

**Figure 1:**
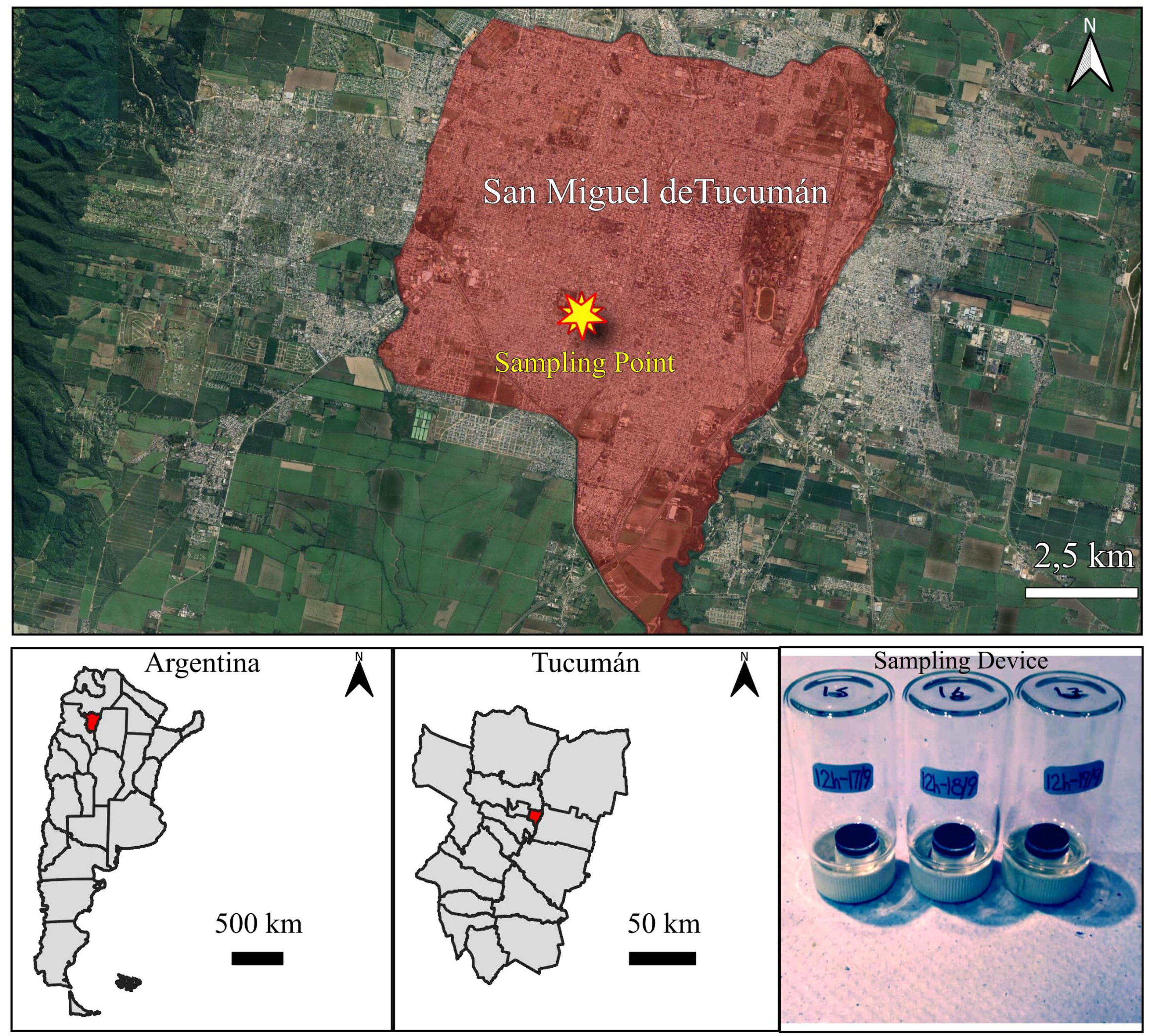

Sampling occurred weekly from August 2019 to August 2020, between 07:00 and 19:00. Rainy days were avoided to prevent interference from precipitation. Sampling paused in January (university recess) and mid-March to mid-April due to the COVID-19 lockdown. Preliminary tests showed that the adhesive surface was effective for up to 36 hours of exposure.

### 2.2. PM Characterization Using SEM

Samples were imaged using a Zeiss Supra 55VP microscope at the Centro Integral de Microscopía Electrónica (CIME, CONICET-UNT). SEM provided three-dimensional images of the sample surface with superior depth of field and detail compared to conventional optical microscopy. This enabled detailed imaging and analysis of particles within the micrometer scale.

Images were acquired using backscattered electrons at 15 kV, a working distance of 8 mm, and an aperture of 60 µm. High-resolution images (3072 × 2304 pixels) were captured at 300x magnification, allowing the analysis of particles with diverse diameters.

### 2.3. Chemical Analysis of PM Using EDS

The chemical composition of individual particles was analyzed using an X-ray spectrometer coupled to the SEM. At least 300 particles were randomly selected and analyzed based on grayscale differences in SEM images. EDS spectra were acquired using Oxford INCA EDS, with the electron beam positioned at the particle center for precise compositional data.

### 2.4. Image Processing and Analysis

SEM images were processed using the free-access software ImageJ® (National Institutes of Health, https://imagej.nih.gov/ij/index.html). Approximately 15,000 particles distributed over the year of sampling were peaked in the SEM images and subjected to image analysis. This allows us to estimate PM concentration levels during the sampling period through particle number measure in a standardized number of microphotographs. Images were scaled by setting a pixel-to-distance measure based on the SEM’s magnification settings. Cropping was applied to exclude parameter bars, and normalization adjusted the background grayscale value to a uniform level of 30. Profile values were plotted, taken along transects that included carbon-rich particles, silica-rich particles, and background to observe limit pixel values between particles and background (Figure 2). This method helped to standardize analysis of SEM images for particle characterization and comparison.

**Figure 2:**
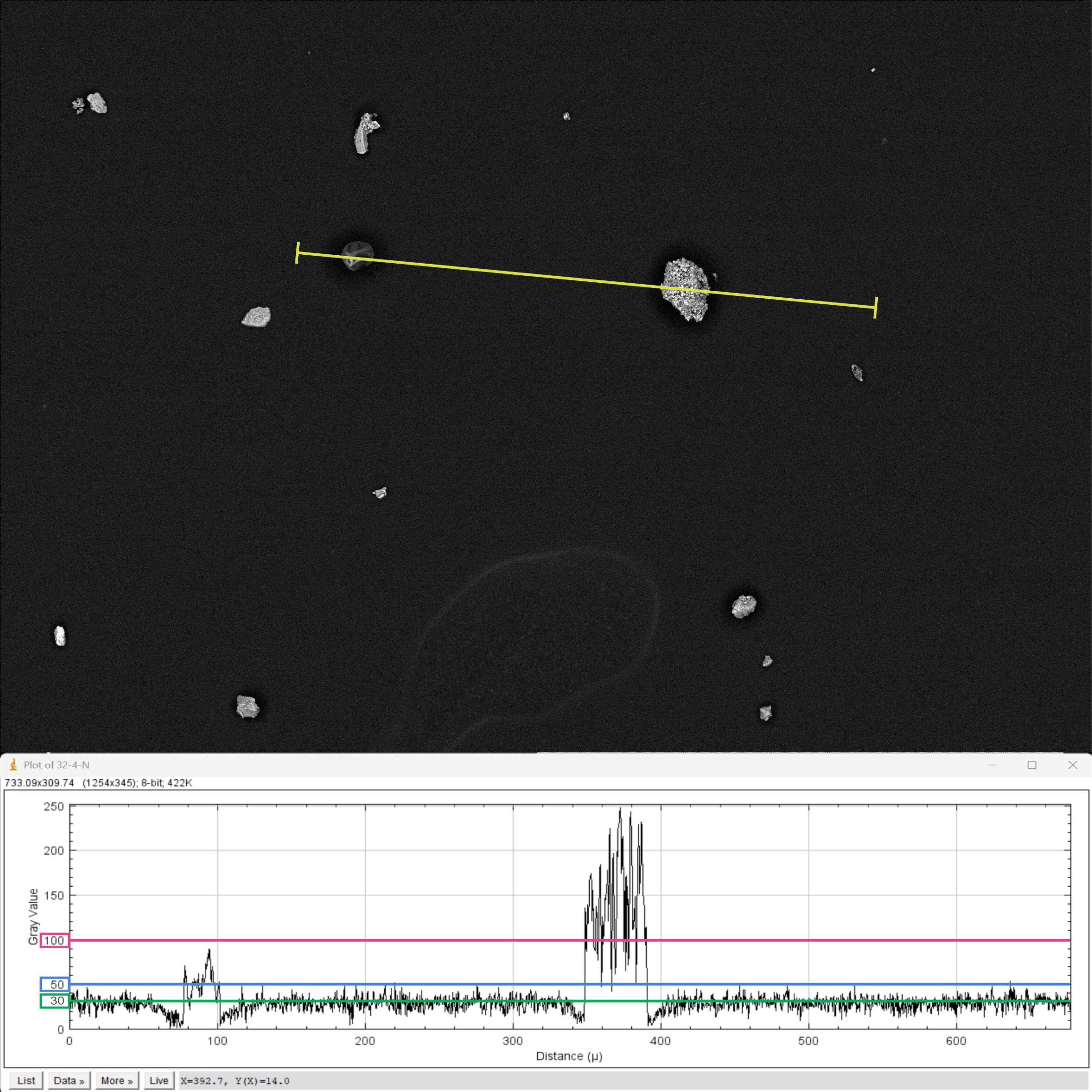

Thresholding was employed to differentiate particles from the background, setting a pixel value range of 50–255 for particles (Figure 3a). Noise and imperfections on the carbon tape were corrected using the “Brush” tool and a median filter. Particle parameters, including area, mean gray value, Feret diameters (maximum and minimum), circularity, and roundness, were measured using the “Analyze Particles” command on the normalized images (Figure 3b).

**Figure 3:**
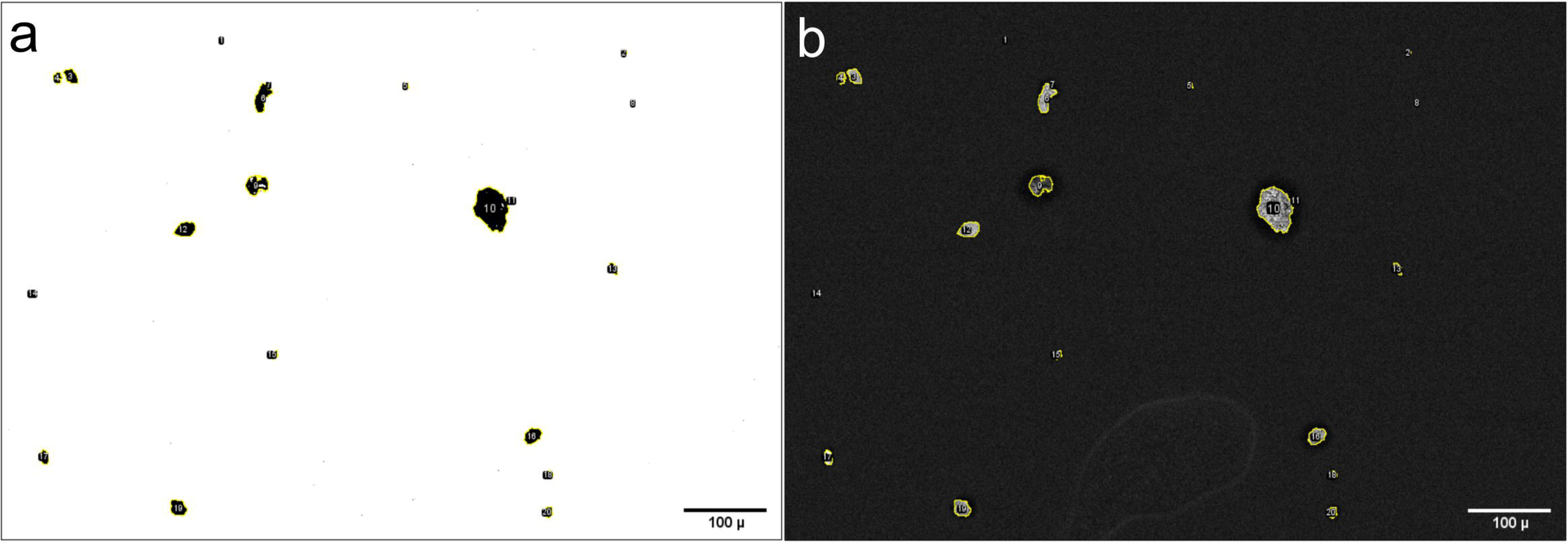

Parameters obtained with ImageJ, such as Feret diameters, were used to calculate the equivalent diameter (D_eq_) of individual particles (Merkus, 2009; Sgrigna et al., 2016). Particles were then classified into three distinct categories based on D_eq_: I) Dust: Particles >10 µm; ii) PM_10_: Particles <10 µm and >2.5 µm; PM_2.5_: Particles <2.5 µm.

Ultra-fine particles, such as PM_1_ (<1 µm), were excluded from the analysis for two reasons: i) these particles remain suspended in the air and are less likely to settle on the adhesive surface unless direct contact occurs, and ii) their small size results in higher measurement errors, even with optimal resolution.

For each sample, 12 images were analyzed, and the resulting data were unified using the statistical programming language “R” (https://www.r-project.org/) to facilitate comparison across samples.

### 2.5. Environmental Parameter Analysis

Meteorological data, including temperature, relative humidity, and wind speed, were obtained from the National Meteorological System of Argentina (https://www.argentina.gob.ar/smn). Correlations between these parameters and PM distribution were assessed using Spearman correlation coefficients (*ρ*) calculated with R, which provided insights into the direction and intensity of these relationships.

### 2.6. Nitrogen oxides mapping

For comparison with PM data, concentration and distribution maps of nitrogen oxides (NO-) were generated using QGIS 3.34 software (https://qgis.org/project/visual-changelogs/visualchangelog334/). Sentinel-5P satellite products were obtained from the Sentinel Hub repository via the EO Browser platform (https://apps.sentinel-hub.com/eo-browser). The selected products had been pre-processed by the provider for nitrogen dioxide (NO-), using the S5P/TROPOMI NO- version 2.2.0 dataset, which offers improved sensitivity to these gases and reports their tropospheric column density in mol/m^2^. Only satellite images corresponding to dates without cloud interference were selected. For both 2019 and 2020, the days with the highest and lowest PMC_2.5_ concentrations during the sugarcane harvest season were analyzed, only if the corresponding satellite data were available. For spatial analysis, a study area with a 20 km radius centered on the sampling site was used, extending radially to the boundaries of San Miguel de Tucumán.

## 3. Results

### 3.1. Quantification of PM and Size Distribution Using SEM Image Analysis

SEM image analysis revealed seasonal patterns in PM levels throughout the 2019–2020 period (Supplementary File available at <u>10.5281/zenodo.15520804</u>). Peaks occurred in August and September, during sugarcane harvest, likely due to biomass burning article counts decreased during wetter and warmer months (December to March), confirming the influence of climatic conditions. The COVID-19 lockdown (March to November 2020) also affected PM levels.

During harvest season, PM concentrations increased by 110% in 2019 and 163% in 2020, compared to non-harvest months. Average PM counts were 695 particles/sample in 2019 and 870 particles/sample in 2020, versus 331 particles/sample outside harvest periods (Figure 4). This aligns well with findings from industrial cities, where PM_10_ and PM_2.5_ concentrations were found to be up to 2.0 and 1.7 times higher than national standards due to industrial and vehicular emissions (Xu et al., 2019).

**Figure 4:**
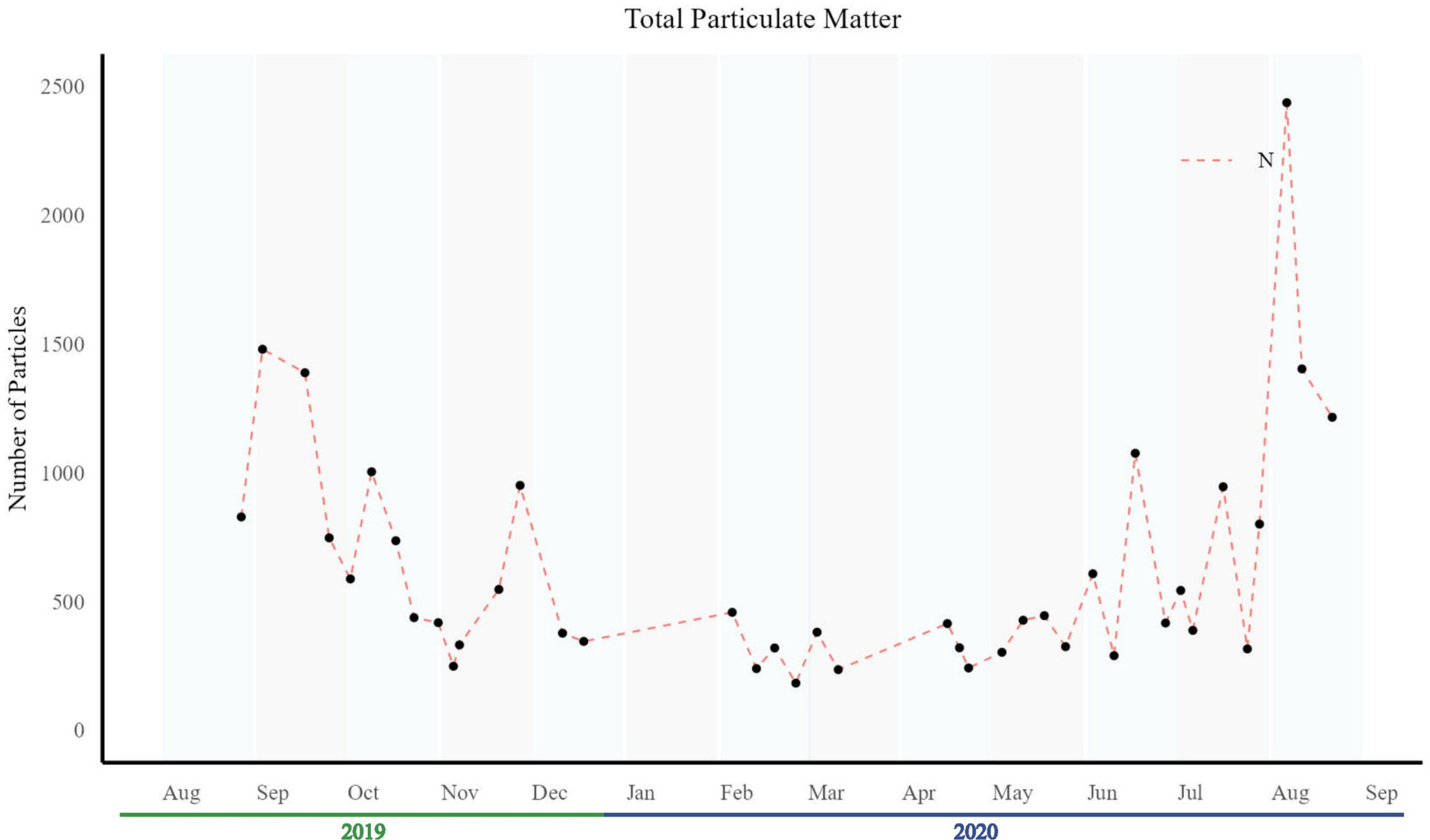

Dust, PM_10_, and PM_2.5_ levels were highest during harvest season (Figure 5). In 2019, finer particles (PM_10_ and PM_2.5_) dominated, suggesting combustion sources. In contrast, in 2020, all size fractions increased more uniformly, highlighting the influence of multiple PM sources and meteorological effects on particle distribution.

**Figure 5:**
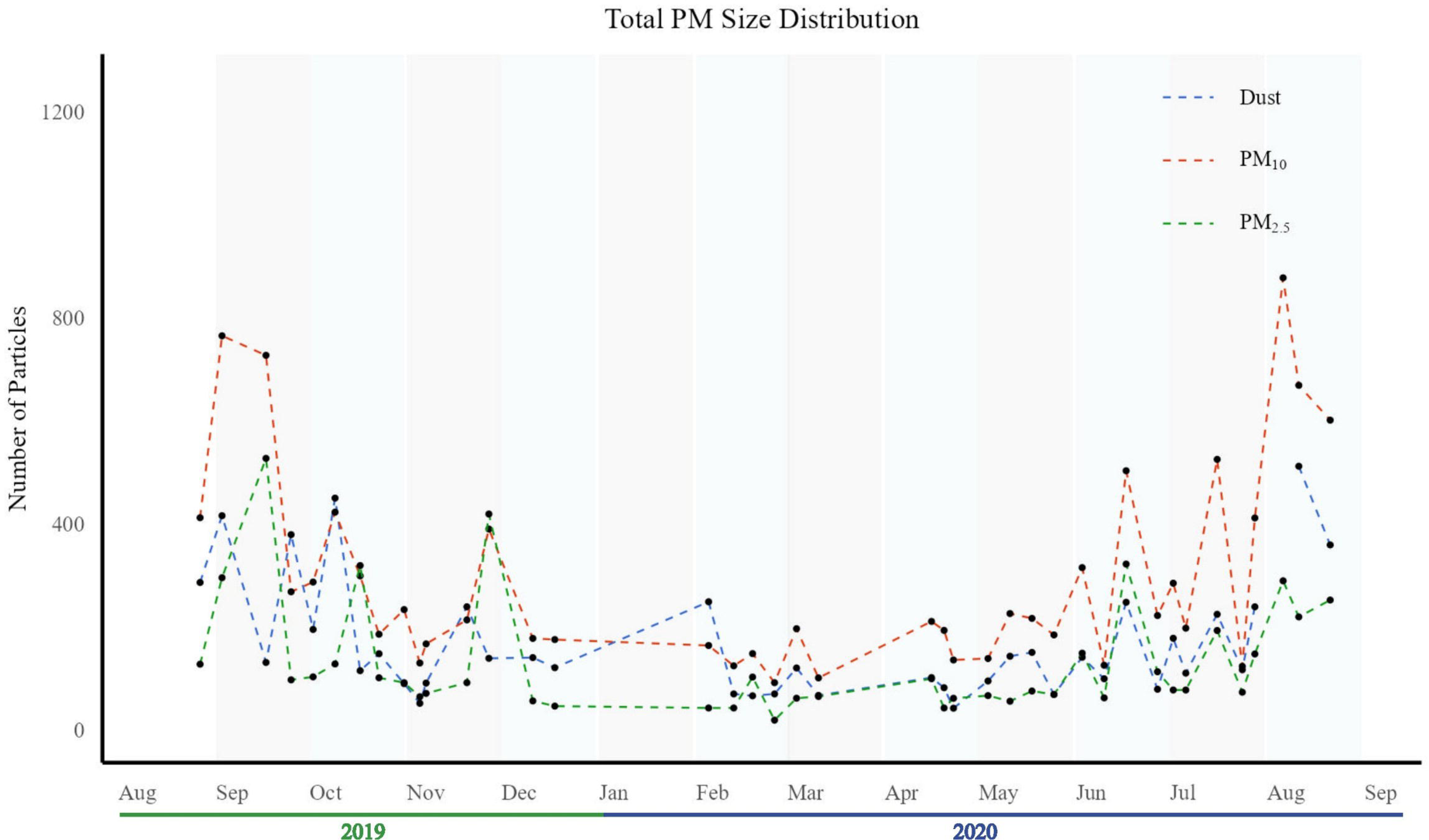

### 3.2. Chemical Composition and Classification of PM

EDS analysis identified consistent elemental composition across the sampled particles, with predominant elements being carbon (54.3%), silicon (26.13%), aluminum (4.82%), potassium (3.90%), calcium (2.73%), and iron (2.82%). This enabled classification into two main groups: i) Carbonaceous Particulate Matter (PMC): predominantly composed of carbon, associated with “black carbon” or “total carbon” from biomass burning and combustion (Shen et al., 2021); and ii) Terrigenous Particulate Matter (PMT): composed primarily of silicon, aluminum, potassium, calcium, and iron, characteristic of “soil dust” or “mineral dust,” typically derived from rock and soil-forming minerals(Camino *et al*., 2015; Hosseini and Shahbazi, 2016).

The mean gray value of SEM images served as an indicator of chemical composition, enabling the discrimination between PMC (gray values of 30–100) and PMT (gray values >100). This approach facilitated the analysis of particle distribution and compositional variability, revealing a clear dominance of PMT over PMC throughout the year (Figure 6a and 6b), except in PM_2.5_, fraction, which exhibited a high concentration on PMC (Figure 6c). Compared with the non-harvest season there is an average increase of 92% for DustT, 103% for PMT_10_ and 211% for PMT_2.5_ during harvest season of 2019, and 207% for DustT, 171% for PMT_10_ and 253% for PMT_2.5_ during harvest season of 2020. A similar pattern was observed for PMC, the increase is 119% for DustC, 70% for PMC_10_ and 144% for PMC_2.5_ during harvest season of 2019, and 49% for DustC, 42% for PMC_10_ and 103% for PMC_2.5_ during harvest season of 2020.PMTwas more abundant, likely due to drier soil conditions and wind erosion, suggesting that environmental conditions could play a crucial role in PM size composition and distribution (Yadav et al., 2017). The high content of PMC in finer particles could be explained by increased combustion activity, either from biomass burning or fossil fuel combustion, as reflected in PMC_2.5_ levels (Grange et al., 2022; Paisi et al., 2024).

**Figure 6:**
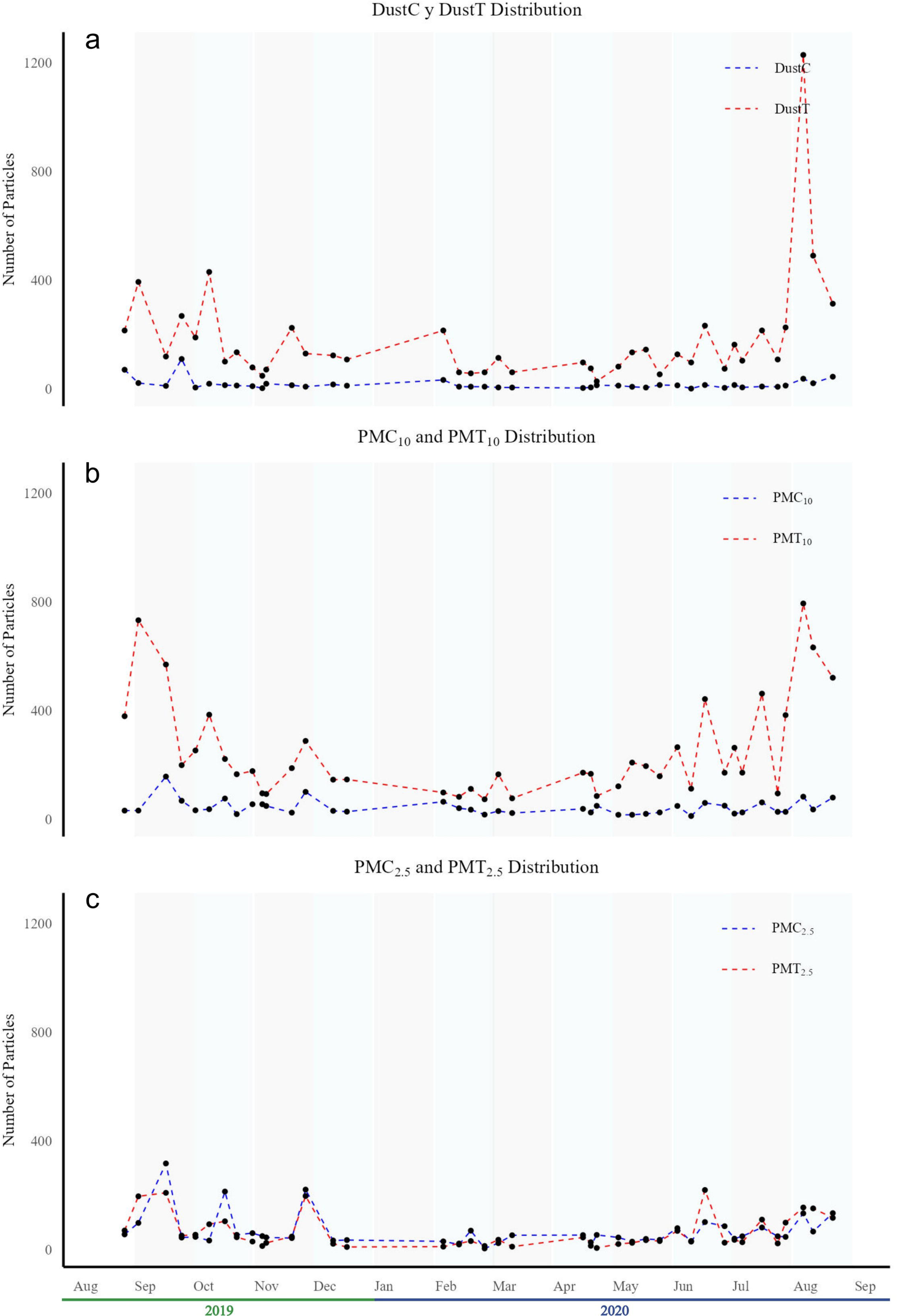

### 3.3. Environmental Influences on PM

Environmental parameters such as humidity, wind speed, and temperature influenced PM transport and deposition (Figure 7), as evidenced by the Spearman correlation coefficient (*ρ*). In this case, humidity was negatively correlated with PM (*ρ*=−0.572), indicating particle settling during high humidity. Likewise, wind speed showed a positive correlation (*ρ*=0.398), suggesting the resuspension of particles from surrounding surfaces under windy conditions. Lastly, temperature exhibited a negative correlation (*ρ*=−0.386), potentially reflecting increased particle deposition during periods of high humidity and temperature.

**Figure 7:**
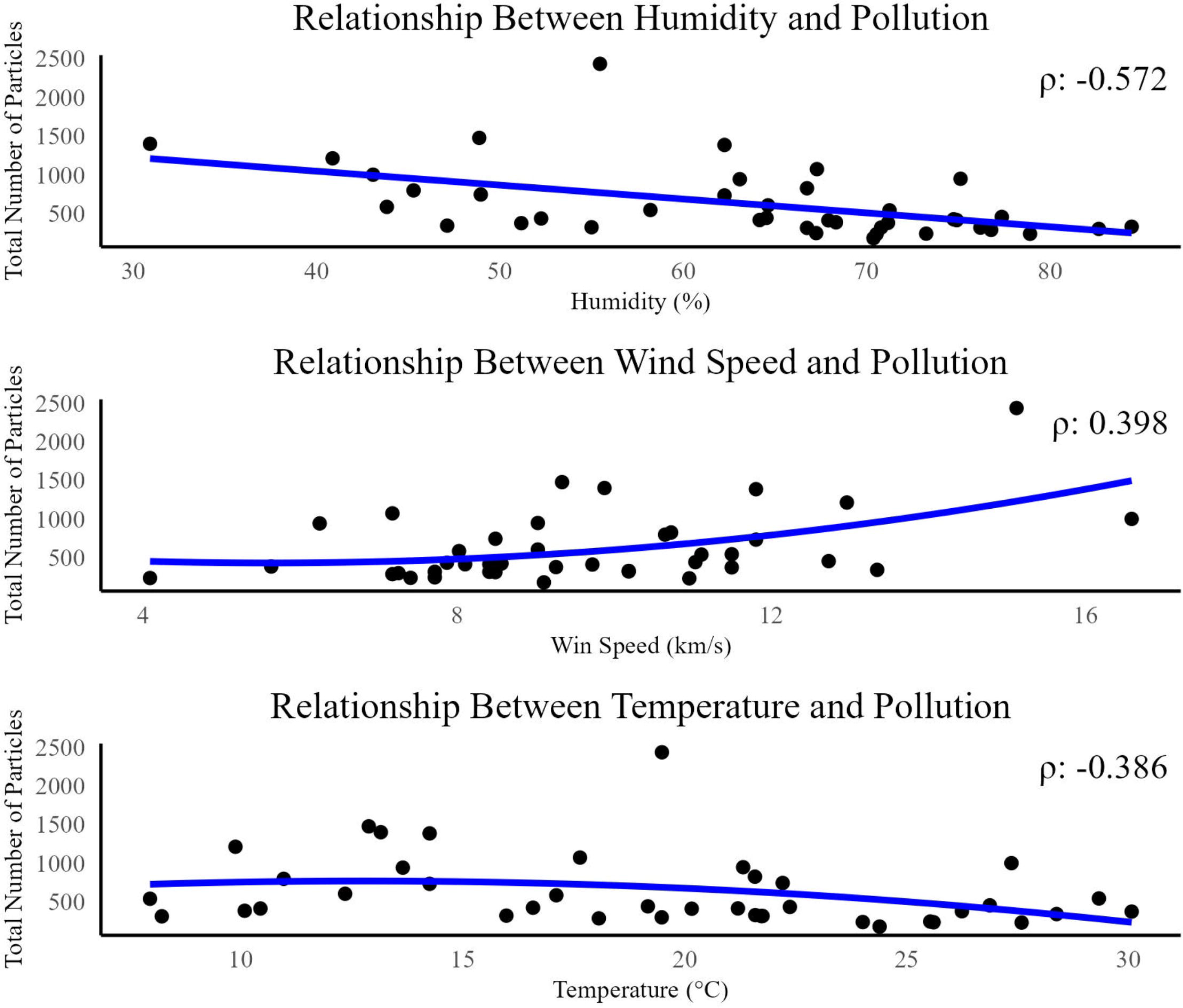

These findings highlight the importance of meteorological factors in PM dynamics. Wind erosion appears to be a significant driver of PMT levels, while humidity plays a key role in reducing airborne PM through wet deposition. That is why during dry season, coinciding with harvest period, the presence of PMT in the air of San Miguel de Tucumán is higher than wet or non-harvest season. This behavior is similar to that observed by Yadav et al. (2017,) described in India during monsoon season, and Zalakeviciute et al. (2020) in Ecuador during the transition between dry and wet seasons.

### 3.4. Sugarcane Burning and PMC Contributions

The anthropogenic activities also contribute to the presence of PM in the air of San Miguel de Tucumán. Finer PMC generally comes from combustion processes like biomass burning and fossil fuel combustion (Grange et al., 2022; Paisi et al., 2024). In this case the most relevant source of biomass burning, originates from the important industrial activity in Tucumán province, the sugar industry (García et al., 2018). Sugar cane fields burning is a significant contributor to PMC_2.5_ to the air. As mentioned in section 3.3, the PMC increase during sugarcane harvest season but the size range more abundant is the PMC_2.5_, with an average ratio PMC_2.5_:PMT_2.5_ of 1:0.9 in the year 2019 and 1:1.2 in the year 2020 (Figure 6c).This would indicate that there was more contribution of biomass burning during the year 2019 than the year 2020, but according to the survey of burned areas in the productive area of the province of Tucumán during the 2020 crop season done by the Estacion Experimental Agroindustrial Obispo Colombres (EEAOC, 2020), there were more surface burned during 2020, 61.000 ha more than 2019. This shows that even though biomass burning is an important source of PMC_2.5_, there is another one to be considered: the fossil fuel combustion associated with vehicle fleet, it was providing more particles in 2019 than 2020.

### 3.5. Influence of Population Mobility and NOx

The decrease of PMC_2.5_ from 2019 to 2020 is interpreted as a direct result diminution of combustion vehicles circulating around the sampling zone and in all San Miguel de Tucumán, due to COVID-19 lock down, during the year 2020. One of the main contributions of fossil fuel combustion to the air is fine PMC_2.5_ (Grange et al., 2022; Paisi et al., 2024). The lockdown significantly reduced population mobility, as shown in the Google Mobility Report (https://www.google.com/covid19/mobility/?hl=es) for the region of San Miguel de Tucumán, therefore it also reduced PMC_2.5_.

To support this hypothesis, data from satellite-derived nitrogen oxides (NO_x_) distribution maps were used (Figure 8). NO_x_ is considered as one of the criteria pollutants according to the United States Environmental Protection Agency (EPA). Includes gases such as NO_2_ and NO, highly reactive gases that primarily come from the burning of fuel. The analysis made considered dates where peaks in PMC_2.5_ concentration were observed in the study area as well as days with lower PMC_2.5_ concentrations. The distribution of NO_x_ gases shows that concentration of this gas was high during the peak of PMC_2.5_ in 2019 (Figure 8a), while it decreased on dates when PMC_2.5_ was lower (Figure 8b). In 2020, this pattern remained consistent for both pollutants (Figure 8c and 8d); however, when comparing the two years, it is observed that pollution levels in 2020 were lower than in 2019. These results show that NO_x_ concentration decreased in 2020, corresponding to the period when both population mobility and PMC_2.5_ concentration declined. Additionally, the NO_x_ concentration observed in 2019 could be considered a reference or ‘normal’ value for a year without confinement measures, which again aligns with the high PMC_2.5_ concentration, supporting the role of traffic emissions in PMC_2.5_ presence.

**Figure 8:**
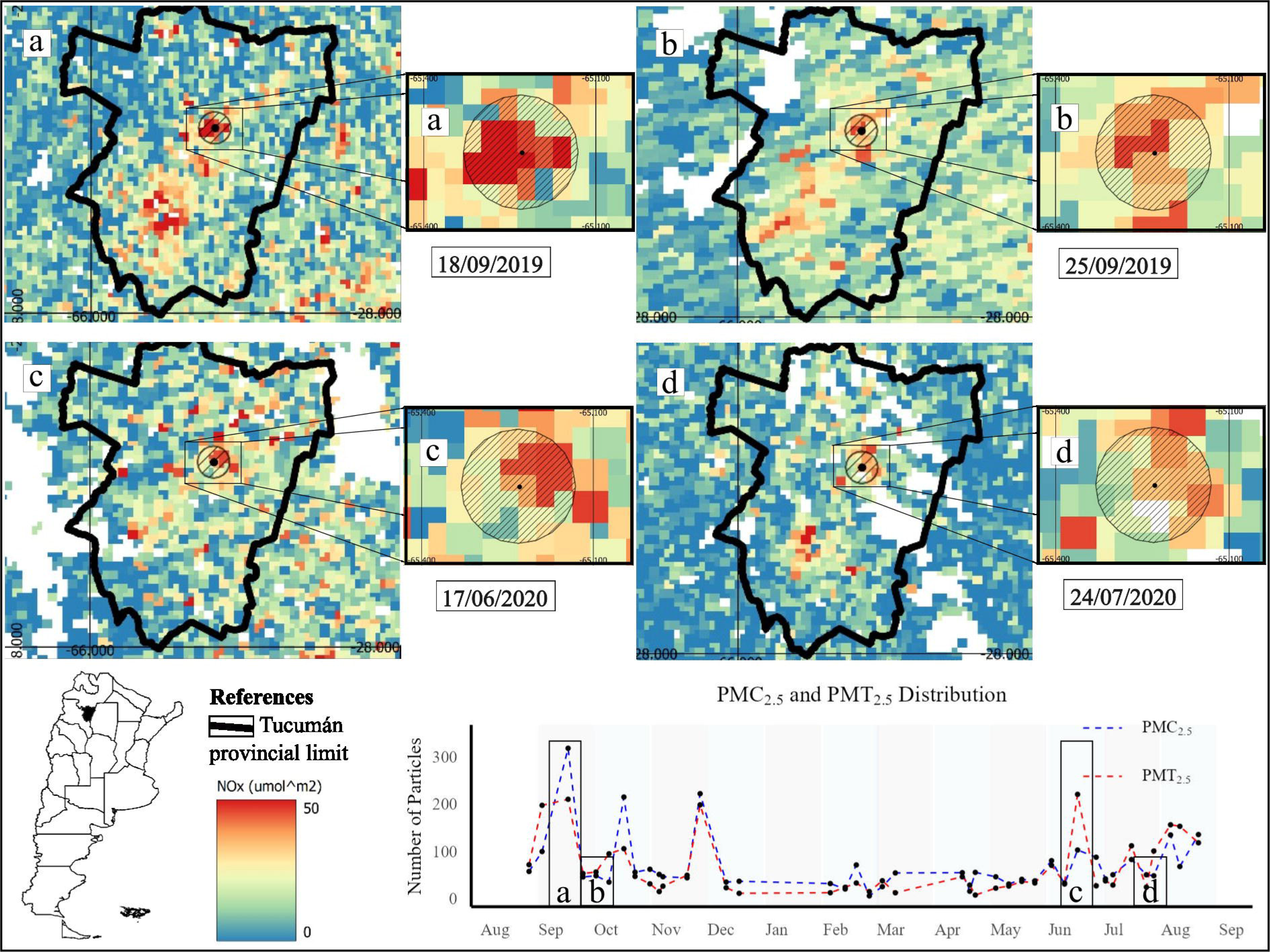

## 4. Discussion

The results of this study highlight the complex interplay between seasonal agricultural practices, industrial activity, urban mobility, and atmospheric conditions in shaping air quality in San Miguel de Tucumán. The increase in PM_2.5_ concentrations during the sugarcane harvest season aligns with previous research linking biomass burning to elevated fine particulate levels in South American cities (Mendez-Espinosa et al., 2019). However, the observed decline in PMC_2.5_ in 2020 -despite a larger burned area-suggests that vehicular traffic contributes more substantially to carbonaceous fine particles than previously acknowledged. This is consistent with findings from urban environments in Europe and Asia, where traffic emissions dominate PM_2.5_ sources (Grange et al., 2022; Tessum et al., 2022; Liao et al., 2023; Paisi et al., 2024). This is further supported by the positive correlation between NOx satellite data and PMC_2.5_ levels, reinforcing the role of human mobility in combustion-related emissions.

These insights support the growing consensus that integrated monitoring strategies—including high-resolution morphological and chemical analyses—are essential for understanding urban air pollution dynamics and designing targeted mitigation policies (Baldacchini et al., 2019; Chen et al., 2022). In this context, SEM-EDS emerged as a scalable and precise tool for PM source apportionment, especially in mid-sized cities where conventional monitoring networks are limited.

By analyzing over 15,000 particles collected in one year, we identified significant temporal and compositional variability in PM related to anthropogenic activities such as sugarcane burning and traffic, modulated by meteorological factors. SEM-EDS enabled the classification of particles into carbonaceous (PMC) and terrigenous (PMT) fractions, providing insights into seasonal emission sources. Its capacity to integrate morphology and elemental composition at the microscale proved critical for source attribution under complex atmospheric conditions (Martínez Morales et al., 2023; Pandey and Negi, 2022).

The COVID-19 lockdown offered a unique opportunity to disentangle the effects of reduced human activity. Our results revealed a strong association between reduced traffic and lower PMC_2.5_ levels, consistent with findings from other regions where mobility restrictions led to significant decreases in PM_2.5_ levels (Arregocés et al., 2021; Martínez Morales et al., 2023). However, other studies have highlighted that such reductions are not always straightforward. For instance, Zangari et al. (2020) reported that changes in PM levels during the lockdown period were within the range of interannual variability, suggesting that natural variability and meteorological conditions also play a role. Pandey and Negi (2022) found that biomass burning was the primary contributor to PM during lockdowns in their study area, underscoring the regional specificity of emission sources. Similarly, Speranza and Caggiano (2022) further noted that pandemic-related mobility restrictions affected both coarse and fine PM fractions differently across study sites, indicating that the impact of lockdowns on PM composition is spatially heterogeneous. Post-pandemic analyses, such as Qonitan et al. (2023) in Jakarta, Indonesia, showed a deterioration in air quality linked to vehicular and coal combustion emissions. Taken together, these findings reinforce the value of SEM-EDS as a diagnostic tool for particle classification and for understanding the multifactorial nature of air pollution, especially under abrupt changes in human activity such as those caused by the COVID-19 pandemic.

A major advantage of SEM-EDS lies in its ability to bridge the gap between high-precision laboratory techniques and real-time monitoring, offering a benchmark for calibrating low-cost air quality sensors. The method’s ability to combine morphological and chemical information enhances its relevance in urban settings with complex emission profiles.

Ultimately, the findings of this study not only demonstrate the potential of SEM-EDS in air quality research but also highlight critical intervention points for environmental policy. In particular, the seasonal impact of agricultural burning and vehicular emissions on urban PM levels urges the need for stricter enforcement of burning bans, improved traffic regulation, and targeted air pollution mitigation strategies in Tucumán and other regions facing similar challenges. Most importantly, our objective goes beyond characterizing air quality: we aim to promote this issue as an urgent public health and environmental challenge. By providing high-resolution data on pollution sources, we seek to empower evidence-based policymaking and emphasize the necessity of strengthening local and regional frameworks to ensure more effective public health protection and long-term sustainable urban development.

## Data Availability

All data produced are available online at

https://zenodo.org/records/15520804

## Credit authorship contribution statement

**Enzo Rubén Marcial:** Conceptualization, Investigation, Methodology, Formal analysis, Validation, Visualization, Writing – original draft. **Alexander Aldo Santucho Cainzo:** Methodology. **Facundo Reynoso Posse:** Investigation, Visualization. **Rodrigo Gastón Gibilisco:** Review& editing; **Aida Ben Altabef**: Review & editing. **Diego Hernando Corregidor Carrió:** Formal Analysis, Investigation, Resources, Supervision, Project administration, review & editing. **Virginia Helena Albarracín:** Funding acquisition, Investigation, Resources, Supervision, Project administration, Writing – review & editing.

## Acknowledgments

The authors acknowledge the generous financial support by the National Council of Research and Technology (CONICET) and ex Ministry of Science, Technology and Innovation of Argentina (MINCyT) through “Programa de Articulación y Fortalecimiento Federal de las Capacidades en Ciencia y Tecnología COVID-19”. This work was funding by the Project TUC-03-COVID19 “COVID-U-MAP”, included in the Bank of Technological and Social Development Projects of CONICET (PDTS). Our group was part of the “Federal Network for COVID detection in the environment”, Unidad Coronavirus, MINCyT. The Project received financial support of the Provincial Health Ministry in the frame of COVID-19 Research Program, processed by a specific agreement between the Superior Government of the Province of Tucumán, the National University of Tucumán and CONICET. ERM is a postdoctoral researcher from CONICET. VHA, RGG, ABA are staff researchers and DHCC and staff technicians. VHA was a recipient of a Georg Foster Scholarship for Experienced Researchers, Alexander von Humboldt Foundation (2021-2023) and of a grant of the Williams Foundation (Fondos Complementarios para Proyectos con Impacto en el Territorio 2024). We would also like to thank technical assistance of Luciano Martinez and Hernán Esquivel for imaging samples. All micrographs and EDS data were taken at the Centro Integral de Microscopía Electrónica (CIME), belonging to UNT and CONICET, in Tucumán, Argentina.

